# Effect of a single dose of oral iron on pancreatic beta-cell function in healthy individuals: a before-and-after (pre-post) study

**DOI:** 10.1101/2022.02.11.22270879

**Authors:** Padmanaban Venkatesan, Jagadish Ramaswamy, S Vanitha, Molly Jacob, Joe Varghese

## Abstract

**Introduction:** Although *in vitro* and animal studies have shown that iron loading in pancreatic beta-cells impaired insulin secretion, no human studies have documented the acute effects of oral iron on beta-cell insulin secretory capacity. In this study, we determined beta-cell insulin secretory capacity at baseline and after a single oral dose of iron (ferrous sulphate, 120 mg elemental iron) in healthy male individuals.

**Methods:** Fifteen healthy male volunteers underwent an oral glucose tolerance test (OGTT) to document baseline glucose tolerance and insulin secretion kinetics (baseline OGTT). One week later, the same subjects underwent a second OGTT, two hours after an oral dose of ferrous sulfate (120 mg of elemental iron) (post-iron OGTT). Changes in disposition index, insulin secretion kinetics, glucose tolerance, insulin clearance, and iron-related parameters in serum were determined.

**Results:** Compared to baseline OGTT, the areas under the curve (AUC) for serum iron and transferrin saturation increased by 125% and 118% respectively, in the post-iron OGTT. The disposition index decreased by 20% (p=0.009) and the AUC for glucose concentrations increased by 5.7% (p<0.001) during the post-iron OGTT. The insulin secretion rate was marginally lower during the first hour (−3.5%, p=0.63), but became significantly higher during the second hour (22%, p=0.005) of the post-iron OGTT. Concentrations of glucose, insulin and C-peptide in blood were significantly higher at 120 min of the post-iron OGTT.

**Conclusion:** The decrease in disposition index and glucose tolerance observed after the oral dose of iron points to an acute iron-induced impairment in pancreatic beta-cell insulin secretory capacity.

## Introduction

Oral iron supplementation is the treatment of choice in patients with mild or moderate iron-deficiency anemia. Oral iron is preferred to intravenous administration because it is convenient, effective, cheap and safe (1). There are many different oral iron preparations and most of them contain iron in the ferrous form (ferrous sulphate, ferrous fumarate, ferrous gluconate, ferrous ascorbate etc.). Although it has been shown that all these preparations are equally effective in increasing hemoglobin levels (1), ferrous sulphate, being easily available and cheap, is the most commonly prescribed iron preparation (2).

Iron is absorbed in the duodenum and proximal jejunum. Dietary iron is usually in the ferric form and must be reduced to its ferrous form prior to absorption. This reduction reaction is catalyzed by duodenal ferrireductases (such as duodenal cytochrome b) and is aided by gastric HCl and other reducing substances in the diet, such as vitamin C (ascorbic acid) (3). Administration of iron in the ferrous form (e.g., ferrous sulphate) circumvents this step, thus making it readily bioavailable. Ferrous iron is transported across the luminal membrane of the enterocytes via divalent metal transporter – 1 (DMT-1). Iron is then transported across the basolateral membrane (into blood) by another transporter, ferroportin. Hepcidin, a peptide hormone synthesized and secreted by the liver, binds to and degrades ferroportin, thus reducing intestinal iron absorption (4).

In the blood, iron is transported bound to the plasma protein, transferrin, which binds iron with high affinity. Transferrin is normally saturated to about 30 to 35% of its total iron binding capacity, leaving a large reserve to bind additional iron. In conditions of iron overload, such as in hemochromatosis or in patients with thalassemia, transferrin saturation can increase significantly. When transferrin saturation increases beyond 60%, and especially as it approaches 80%, a small but significant amount of iron circulates in blood that is not bound to transferrin. This fraction, called “labile iron” or non-transferrin bound iron (NTBI), is highly reactive and can cause oxidative tissue damage (5).

NTBI is rapidly cleared from circulation, mainly by hepatocytes. It has been shown that ZIP14 is physiologically the most important transporter that transports NTBI into hepatocytes (6). Recently, it was shown that ZIP14 is also expressed on human pancreatic beta cells and that it may mediate NTBI uptake by these cells (7).

Several *in vitro* and animal studies have shown that iron overload impairs pancreatic beta-cell function (8–10). Patients with hemochromatosis are known to accumulate iron in the beta cells, resulting in diabetes due to decreased insulin secretory capacity (11,12). On the other hand, iron chelation or dietary iron restriction improves insulin secretion in mouse models of diabetes (13). Similarly, iron chelation in patients with thalassemia major also improved insulin secretion (14). These studies prove a strong link between increased iron and impaired beta-cell function.

It has been shown that, following a single dose of ferrous sulphate (containing 60 - 100 mg of elemental iron), transferrin saturation increased rapidly and peaked (at ∼ 80%) 2-4 hours after administration. This was associated with a significant increase in NTBI, which also peaked at 2-4 hours (15–17). Given that oral iron administration increases NTBI in blood and that pancreatic beta cells take up NTBI via ZIP14 (7), we hypothesized that oral iron may lead to increased iron levels in beta-cells, which may then cause impaired insulin secretion. To test this hypothesis, we conducted a quasi-experimental single arm before-and-after study, where pancreatic beta-cell function was determined at baseline and after a single dose of iron (ferrous sulphate, 120 mg elemental iron) in healthy men.

## Methodology

This study was approved by the Institutional Review Board at Christian Medical College, Vellore, India (IRB no.13294 dt.26.08.2020). It conforms to the guidelines of the Declaration of Helsinki (1975 and subsequent amendments). The study was retrospectively registered with clinicaltrials.gov (NCT05238987).

### Participants

Healthy male volunteers were recruited from among the staff of Christian Medical College, Vellore. Men aged 18 to 60 years and of BMI 18.5 to 30 kg/m^2^ were included in the study, after obtaining written informed consent. We excluded those known to have diabetes mellitus/pre-diabetes and those who were diagnosed with diabetes/pre-diabetes at the time of baseline oral glucose tolerance test (OGTT). Other exclusion criteria were current acute / chronic illness at the time of OGTT (baseline or post-intervention), anemia (Hb < 13 g/dL), current or recent (within 1 month before date of recruitment) history of iron supplementation and presence of gastrointestinal disorders that might affect absorption of iron/glucose.

### Study protocol

OGTT was done twice in each participant, with a one-week gap in between. The first OGTT (baseline OGTT) was done to document baseline insulin secretion kinetics. The second OGTT (post-iron OGTT) was done 2 hours after a single oral dose of ferrous sulphate (120 mg of elemental iron). Details are given in the CONSORT flow-diagram (Supplementary figure 1).

The primary outcome of the study was to determine the change in beta-cell insulin secretory capacity, as measured by the disposition index, following oral iron administration. The secondary outcomes were changes in insulin secretion kinetics, glucose tolerance, insulin sensitivity and insulin clearance rates.

### Protocol for the oral glucose tolerance test

For the OGTT, the participant fasted overnight (8-12 hours); the following morning a fasting blood sample (5 mL) was taken at 8 AM. Following this, 75 grams of anhydrous glucose dissolved in 300 mL of water was given. Blood samples were drawn every 30 minutes for 2 hours (30, 60, 90 and 120 minutes) after the glucose load. Glucose, iron, unsaturated iron binding capacity (UIBC), insulin, and C-peptide were estimated in serum from each sample. Transferrin saturation was calculated using values for serum iron and total iron-binding capacity. Serum ferritin and hematological parameters were measured in the baseline fasting sample.

### Estimation of biochemical and hematological parameters

Plasma glucose, serum iron and UIBC were measured on Roche Cobas c702 auto-analyzer (Roche Diagnostics, Switzerland), serum insulin and C-peptide were measured on IMMULITE 2000 XPi system (Siemens GmBH, Germany) and serum ferritin was measured on Atellica Siemens (Siemens GmBH, Germany) in the Department of Clinical Biochemistry at CMC, Vellore. Complete blood counts and other hematological parameters were measured using Sysmex XN 9000 automated hematology analyzer in the Department of Transfusion Medicine and Immunohematology, CMC, Vellore.

### Determination of glucose tolerance, insulin sensitivity, and insulin clearance rates

Glucose tolerance was calculated using the area under the curve (AUC) for glucose levels during the OGTT. Insulin sensitivity was calculated using the Matsuda index (18) and HOMA-IR (19). Insulin clearance rate was determined as described previously (20).

### Insulin secretion kinetics

Insulin secretion rate was determined by deconvolution of C-peptide levels in blood during the OGTT, based on a previously published mathematical model (21).

### Determination of beta-cell insulin secretory capacity

To estimate beta-cell function, insulinogenic index (22) and disposition index were calculated as described previously (23). Briefly, insulinogenic index is calculated as ratio of change in insulin concentration at 30 min of a OGTT to the change in glucose concentration during the same period (22). Disposition index is calculated as the product of the Matsuda index and insulinogenic index (23).

### Statistical analyses

Data were tested for normality; normally distributed variables are represented by mean and standard deviation and skewed variables by median and inter quartile range. All comparisons between baseline and post-iron OGTT were done using Wilcoxon paired rank test or paired t test, as appropriate. A P value of less than 0.05 was considered statistically significant in all cases. All statistical analysis and visualisation were done using the programming language, R.

## Results

All the 15 participants in the study were men aged 25 to 43 years. At baseline, haematological and iron-related parameters were normal (Tables 1 and 2).

**Table 1.** Characteristics of subjects

| Parameter | Mean ± SD / Median [IQR] |
| --- | --- |
| Age (years) | 31.6 ± 5.9 |
| Height (cm) | 174 ± 8.5 |
| Weight (kg) | 74.2 ± 8.2 |
| BMI (kg/m<sup>2</sup>) | 24.3 ± 1.9 |
| Waist circumference (cm) | 88.7 ± 6.6 |
| Hip circumference (cm) | 98.4 ± 5.1 |
| Waist-hip ratio | 0.92 ± 0.04 |
| Systolic BP (mm Hg) | 119.3 ± 9.5 |
| Diastolic BP (mm Hg) | 76.5 ± 6.9 |
| Serum ferritin (ng/mL) | 48.1 [15.9 – 64.4] |

**Table 2:** Haematological parameters

| Parameter | Mean ± SD/<br>Median [IQR] |
| --- | --- |
| RBC (10<sup>6</sup>/μL) | 5.2 ± 0.4 |
| Haemoglobin (g/dL) | 15.2 ± 0.9 |
| Haematocrit (%) | 45.0 ± 3.2 |
| Mean corpuscular volume (fL) | 85.7 [83.7-87.9] |
| Mean corpuscular haemoglobin (pg) | 29.2 ± 2.1 |
| Mean corpuscular haemoglobin concentration (g/dL) | 33.8 ± 1.4 |
| Red cell distribution width-SD (fL ) | 41.8 ± 4.4 |
| Reticulocyte (%) | 1.4 ± 0.3 |
| Reticulocyte haemoglobin (pg) | 32.3 ± 2.3 |
| WBC (10<sup>3</sup>/μL) | 5.89 [4.48-6.97] |

### Serum iron and transferrin saturation

Compared to baseline values, serum iron levels (Figure 1A) and transferrin saturation (Figure 1C) (measured during the OGTT) more than doubled after the dose of iron. The AUC for serum iron (Figure 1B) and transferrin saturation (Figure 1D), during the post-iron OGTT, increased by 125% and 118% respectively, over baseline values. These increases were statistically significant (P<0.001 in both cases).

**Figure 1:**
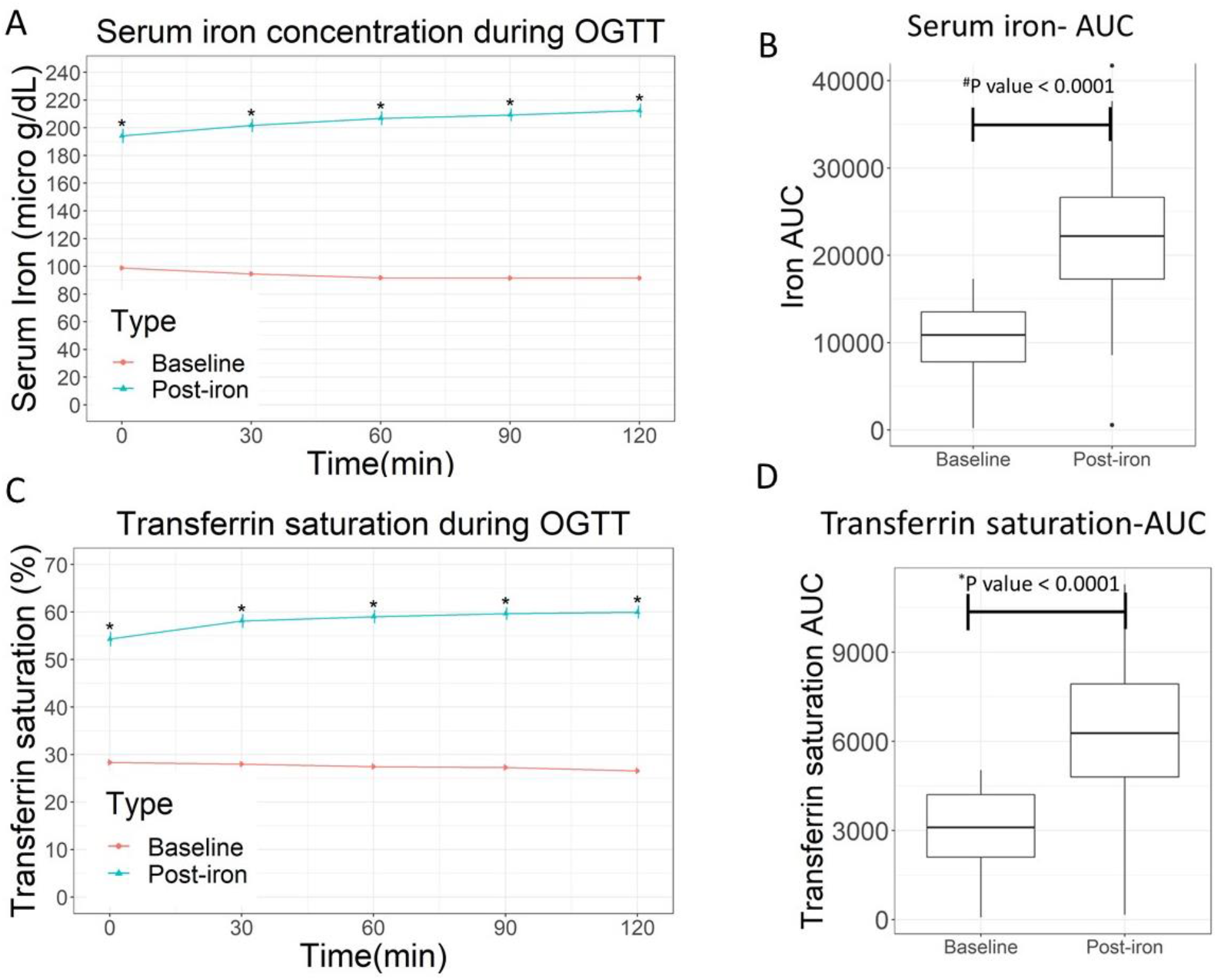
Iron parameters during baseline and post-iron OGTT. *Paired t test P value<0.05, #Wilcoxon ranked test

### Glucose and insulin levels

Glucose values during the first hour of OGTT were similar at baseline and post-iron. However, they were significantly higher at the 90^th^ and 120^th^ minute of post-iron OGTT compared to baseline (P<0.05) (Figure 2A). Overall, the AUC of glucose was 5.7% higher post-iron than at baseline (Figure 2B) (P =0.005).

**Figure 2:**
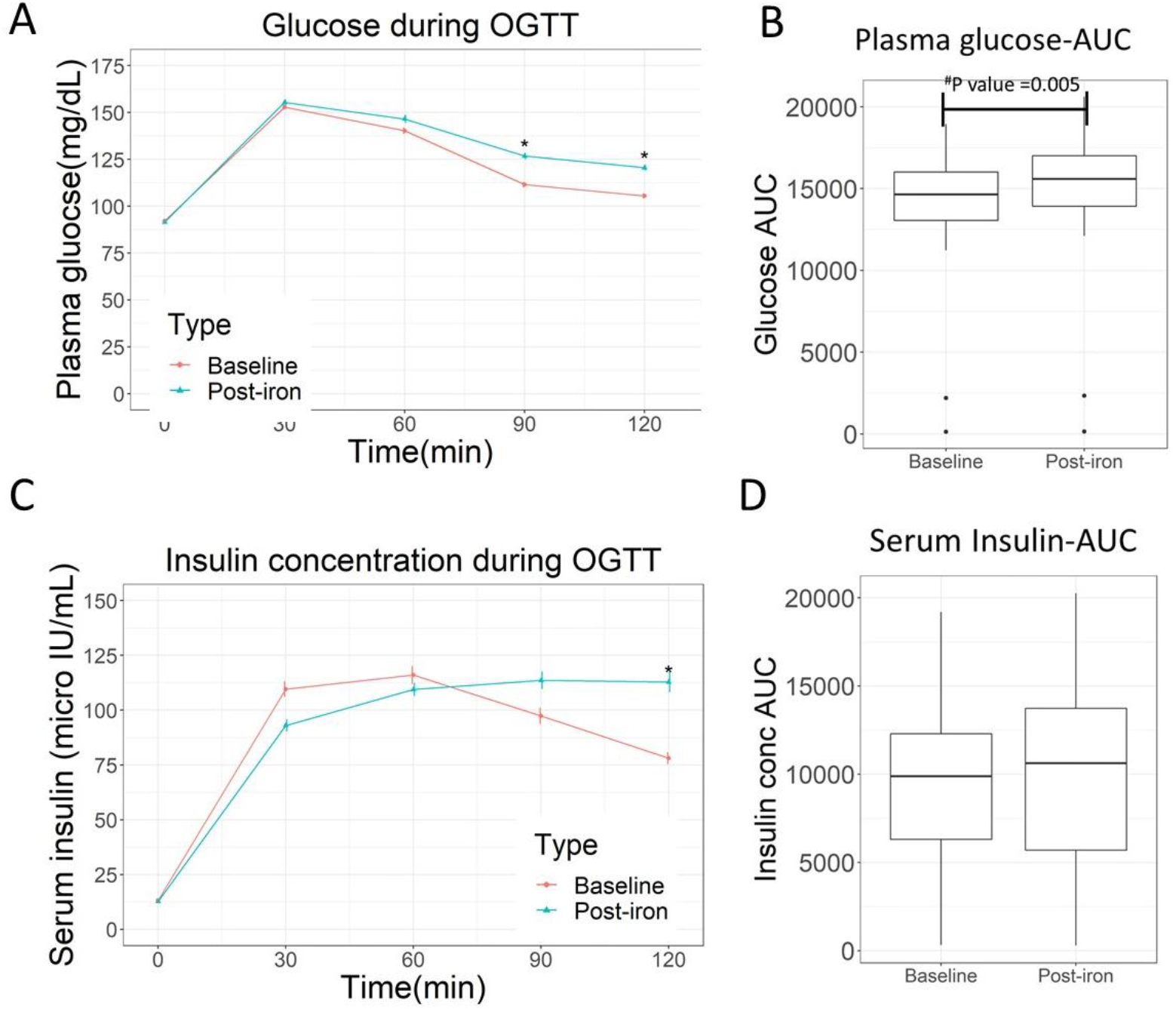
Glucose and insulin concentration during baseline and post-iron OGTT. *Paired T test P value <0.05. # Paired T test

On the other hand, insulin levels were marginally lower during the first hour of the post-iron OGTT, compared to baseline values, but became significantly higher at 120 min (Figure 2C). The AUC for the insulin concentration was, however, not significantly different between the baseline and post-iron OGTTs (Figure 2D).

### Insulin secretion rate

Insulin secretion rate, estimated from C-peptide deconvolution, was similar at baseline and post-iron, during the first hour of the OGTTs. However, it was significantly higher at the last 30 min of the post-iron OGTT, as shown in Figure 3A (P<0.05). Overall, the AUC for insulin secretion was similar during the baseline and post-iron OGTTs (Figure 3B).

**Figure 3:**
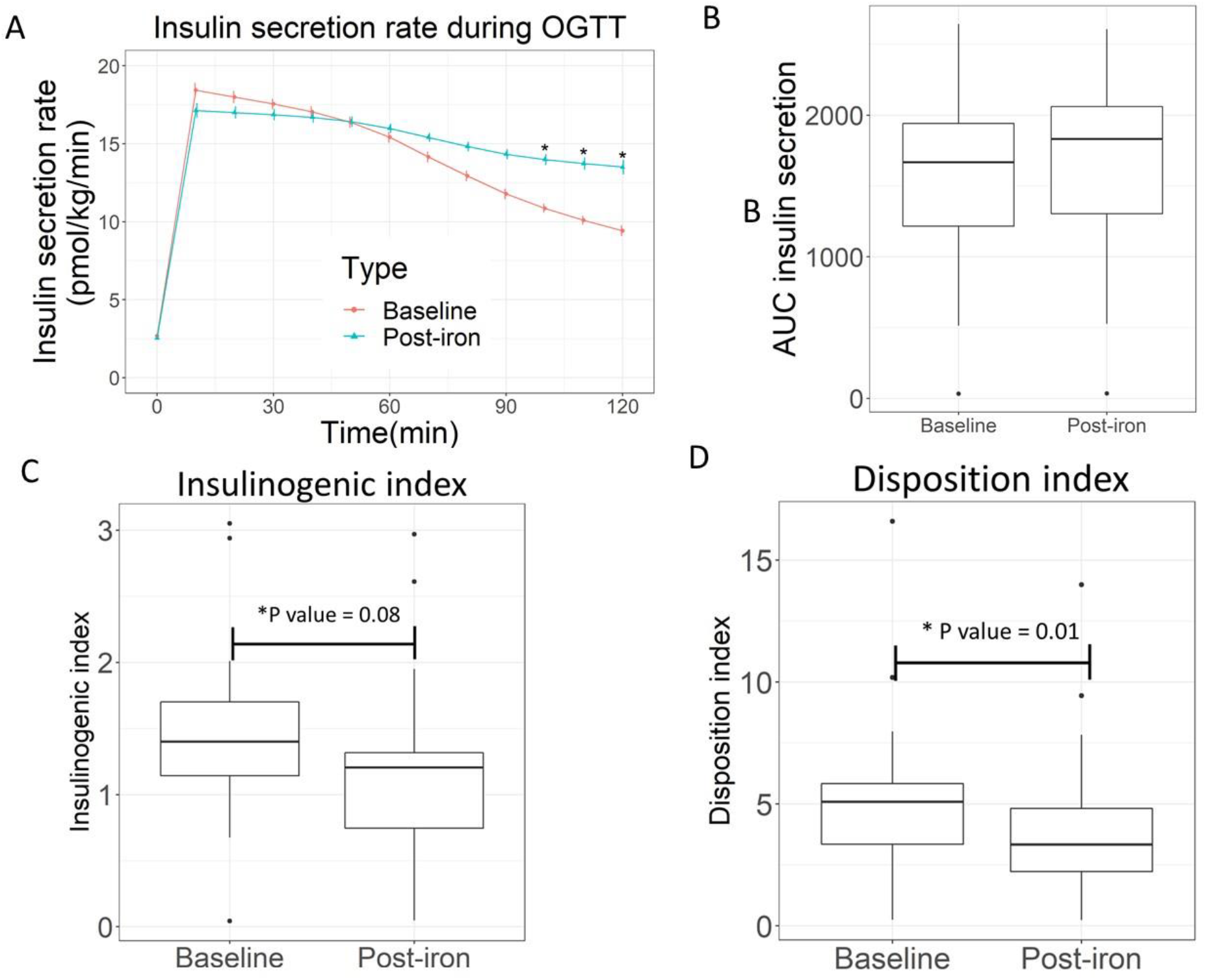
Insulin secretion rate, insulinogenic index, and disposition index. *Wilcoxon ranked test P value <0.05

### Measures of insulin sensitivity and insulin clearance

Insulin sensitivity, as measured by HOMA-IR and Matsuda index, was not significantly affected by the oral iron dose (Supplementary figure 2A and B). The average insulin clearance rates were similar during baseline and post-iron OGTTs (Supplementary figure 2C).

### Measures of beta-cell insulin secretory capacity

Disposition index, an integrated measure of beta-cell compensation in response to changes in insulin sensitivity, was reduced by 20% after the iron dose, compared to the baseline value (P=0.01) (Figure 3C). Insulinogenic index, another measure of beta-cell function, also decreased by 20% post-iron (P = 0.08) (Figure 3D).

### Correlation analysis of changes in transferrin saturation and insulin secretion

To understand the relationship between change in insulin secretion rate and transferrin saturation, we looked for correlations between the change in transferrin saturation and the corresponding change in the insulin secretion rate. There was a statistically significant negative correlation between the two (r = -0.71, P=0.0064).

## Discussion

Studies have shown that disorders with iron overload, such as hereditary hemochromatosis and thalassemia, are associated with development of diabetes mellitus (24–27). Increases in iron stores, even without overt iron overload, have also been reported to be associated with increased risk of developing diabetes mellitus (28–32). While the mechanistic basis of the association between iron stores and diabetes mellitus is still unclear, beta-cell dysfunction is hypothesized to be at least partly responsible for it (33,34).

A mouse model of hemochromatosis (Hfe knockout mice) has shown increased iron accumulation in beta-cells, increased oxidative stress, increased apoptosis of beta-cells and decreased insulin secretion (35). Another study showed that repeated intraperitoneal injection of iron in rats resulted in necrotic changes in beta cells (36). *In vitro* studies, where rat pancreatic islets were incubated with iron, showed increased oxidative stress in and death of beta-cells (37). Exposure of MIN6 cells (a pancreatic beta-cell line) to iron also resulted in increased oxidative stress, decreased cellular insulin content and insulin secretion, and reduced viability of beta-cells (38). Reducing iron levels in diabetic mouse models, through dietary restriction or chelation, resulted in improved insulin secretion (13).

Studies on humans exploring the effect of iron on beta-cell function are scarce. A meta-analysis of epidemiological studies has identified a positive association between dietary intake of heme iron and risk of developing diabetes mellitus (39); however, the study found no increased risk of developing diabetes mellitus among people on iron supplementation (39). On the other hand, studies in pregnant women have found an association between intake of oral iron supplements and incidence of gestational diabetes mellitus (40,41). As discussed in the introduction section, oral iron intake is associated with generation of significant amounts of NTBI in blood (15–17). The fact that beta-cells can take up NTBI via ZIP14 suggests the possibility of acute iron-induced beta-cell damage. To the best of our knowledge, there are no studies that have investigated the acute effect of oral iron supplementation on beta-cell function in humans. To address this, we studied the effect of a single dose of oral ferrous sulphate (120mg elemental iron) on glucose tolerance and insulin secretion in 15 healthy men. We did not recruit women since anemia and decreased body iron stores is common among women which could affect the study results.

Ingestion of 120 mg of elemental iron in the form of ferrous sulphate more than doubled the serum iron levels and transferrin saturation in 2 hours, indicating rapid absorption of iron as would be expected for a highly bioavailable form of iron (42). It resulted in a decrease in glucose tolerance and a reduction in beta-cell function, as measured by disposition and insulinogenic indices.

Measures of insulin sensitivity and insulin clearance rate were not significantly affected by the iron dose. While the decrease in beta-cell function after iron intake is consistent with animal and cell culture studies on the effect of iron on beta-cell function (35,38), it is far from the full picture.

Iron has been shown to produce dual effects on beta-cell function (33,34).While excess iron may result in beta-cell dysfunction, iron in adequate amounts is necessary for the synthesis and secretion of insulin. Iron is an essential component of iron-sulfur clusters of the electron transport chain, and is therefore, required for ATP production and exocytosis of the insulin granules (33). It is also necessary for tRNA modifying enzyme, Cdkal1, which is involved in insulin synthesis (43). More importantly, in the context of acute increases in iron in blood (as in the present study), iron can increase insulin secretion through increased formation of reactive oxygen species, which is known to stimulate exocytosis of insulin granules (33,44,45). On the other hand, excess iron can lead to beta-cell dysfunction, possibly by disruption of the mitochondrial membrane potential, excess production of reactive oxygen species resulting in oxidative stress, impaired synthesis and secretion of insulin, and apoptosis of beta cells (33–35,46).

The observations of the present study are in line with this dual effect of iron on beta-cell function. As seen in Figure 4, subjects with comparatively moderate increases in transferrin saturation (less than 100% increase over baseline) after the iron dose showed improved insulin secretion (compared to baseline). On the other hand, those with greater increases in transferrin saturation (more than 100% increase over baseline and, therefore, possibly greater increases in non-transferrin bound iron) showed lower insulin secretion post-iron, compared to baseline. Since it has been shown earlier that increases in transferrin saturation in response to an oral iron dose was greater in those with low body iron stores (serum ferritin <25 ng/L), compared to those who were iron-replete (serum ferritin >25 ng/L) (17), it is likely the iron-induced beta-cell dysfunction would also be greater in those with low iron stores.

**Figure 4:**
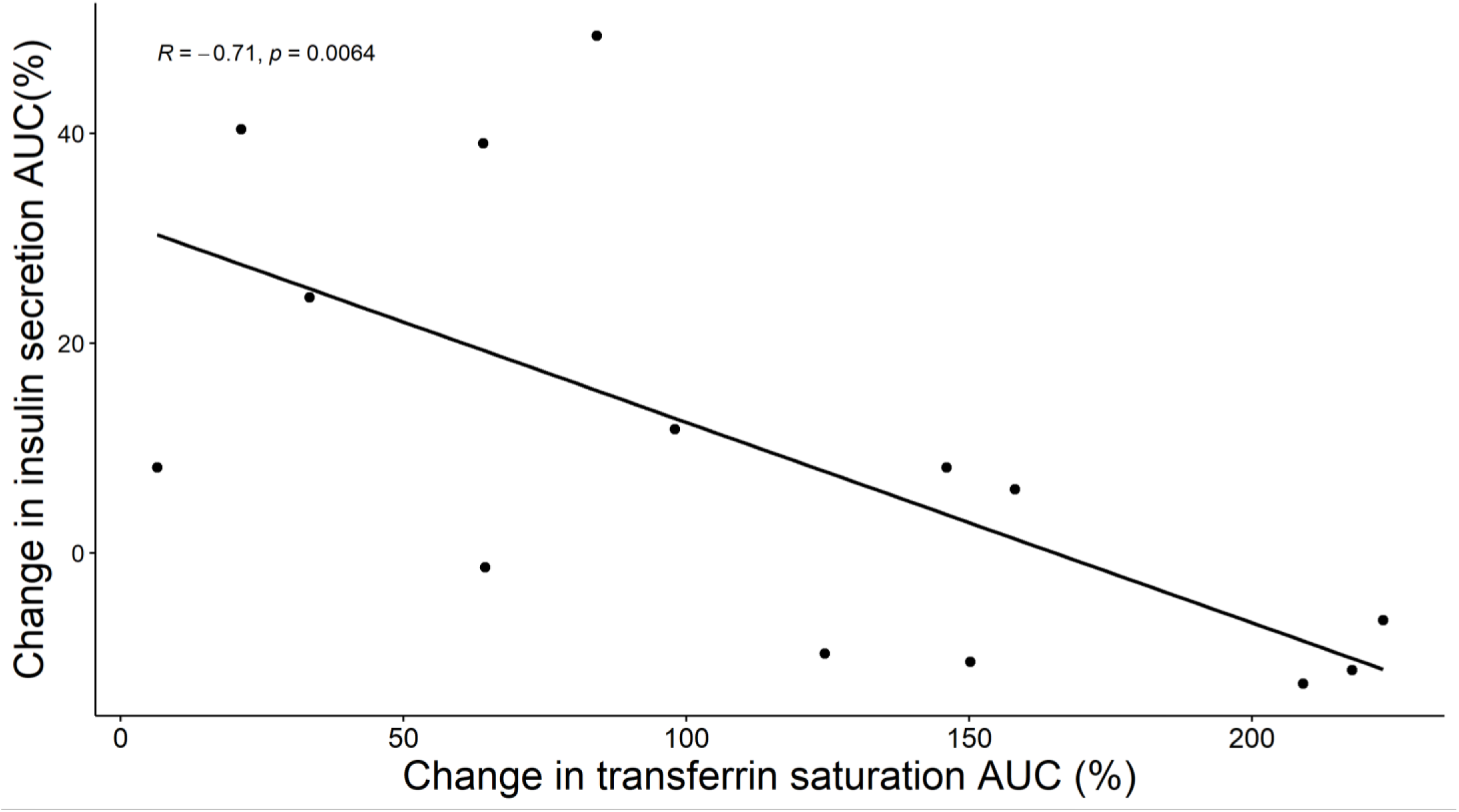
Change in insulin secretion rate and transferrin saturation. An outlier with a transferrin saturation change of 400% is removed from the plot and correlation analysis

Limitations of this study include the small sample size and inclusion of only men. Nevertheless, our findings highlight the deleterious effects of even a single dose of oral iron on beta-cell function. It is not clear if long-term iron supplementation will result in greater beta-cell dysfunction or death. This assumes importance in the context of routine iron supplementation that is commonly recommended in pregnancy women and the risk of gestational diabetes mellitus (40,41). Additional studies are required to investigate this.

In conclusion, we show that a single dose of oral iron acutely lowered glucose tolerance and impaired beta-cell function. Our findings warrant further study of both acute and long-term effect of iron supplements on beta-cell function in clinically relevant populations, such as those with anemia and in pregnant women.

## Data Availability

De-identified data produced in the present study are available upon reasonable request to the corresponding author

## Acknowledgements

We would like to thank the participants of this study.

