## Supplementary figures and images for "Effect of a single dose of oral iron on pancreatic beta-cell function in healthy individuals: a before-and-after (pre-post) study"

### Supplementary figure 1

#### Slide 1
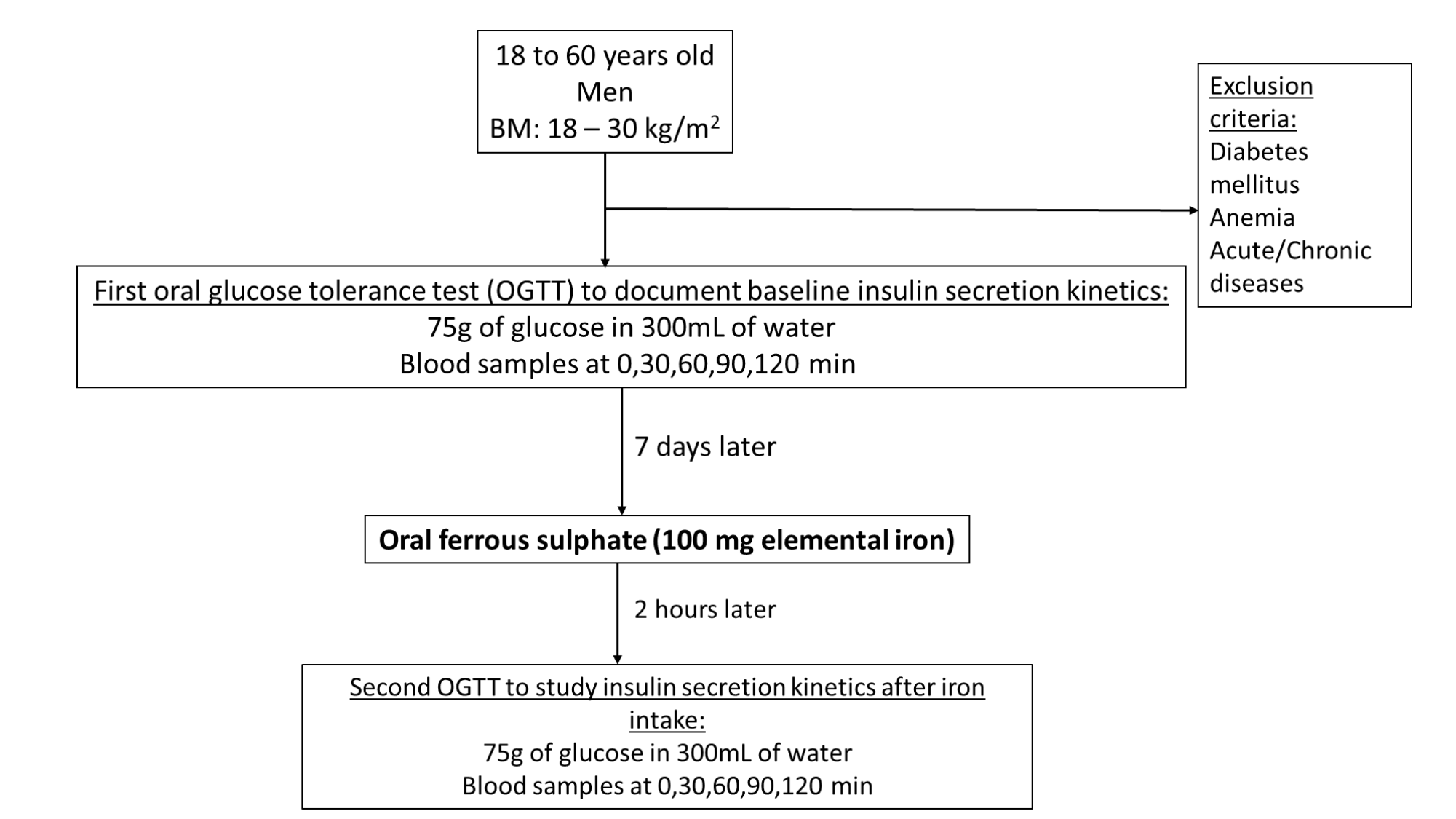

### Supplementary figure 2

#### Slide 1
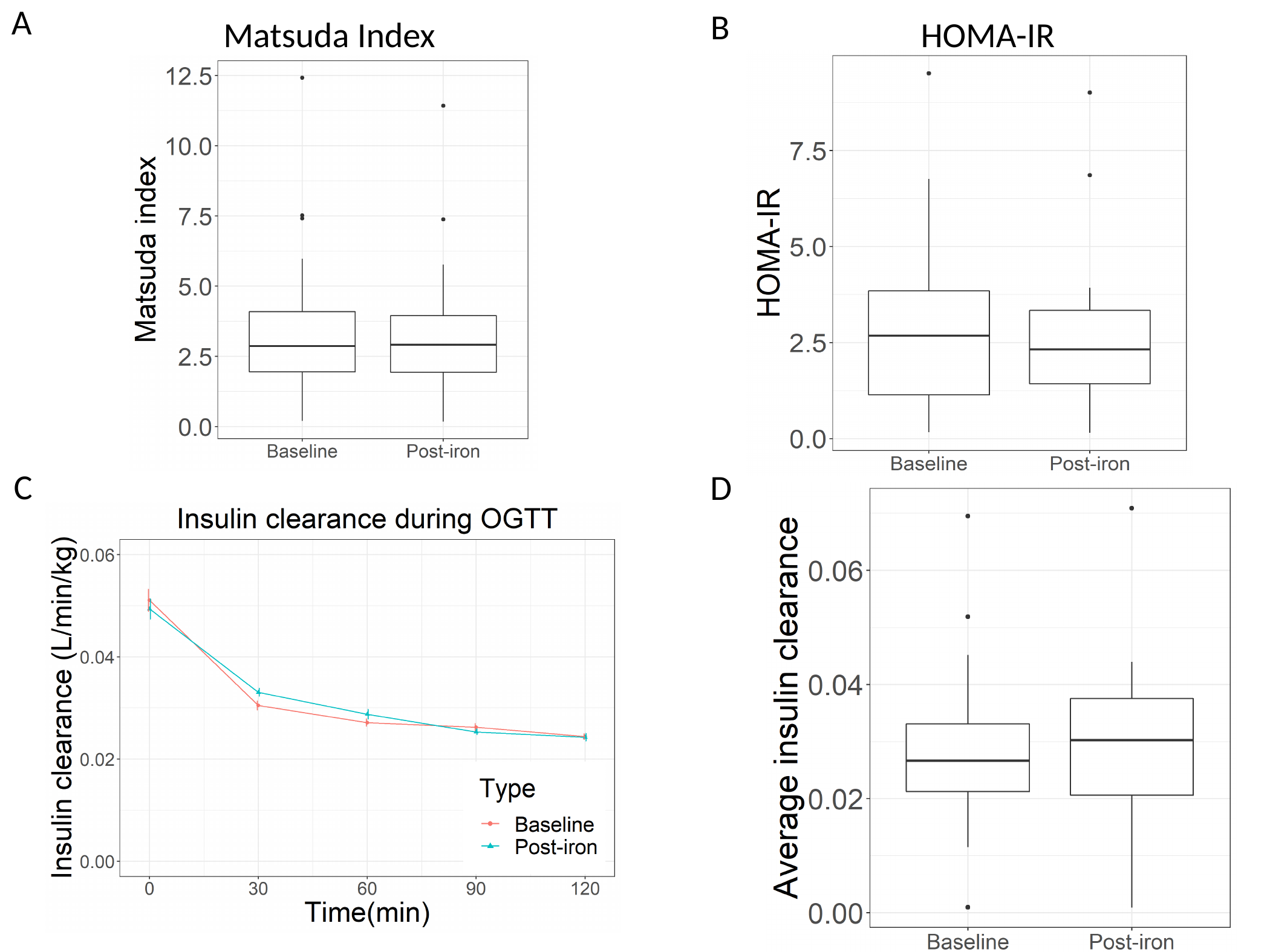

B
A
Matsuda Index
HOMA-IR
C
D
